# Quantifying interocular asymmetry of axial elongation in childhood myopia: metric framework, prevalence, and age-related dynamics

**DOI:** 10.64898/2026.07.18.26358405

**Authors:** Rodion Nesterenko

**Author notes:** **Corresponding author:** Rodion Nesterenko, MD; Eurostyle Clinic, ul. Molodezhnaya 3b, Barnaul 656038, Russian Federation.

## Abstract

**Purpose:** Interocular asymmetry of axial elongation predicts accelerated myopia progression in adults, but childhood prevalence and dynamics are uncharacterised, with no standardised metric. A scale-invariant metric is proposed and characterised in a paediatric cohort with progressive myopia.

**Methods:** A retrospective cohort of 267 children (5–16 years; 913 follow-up intervals) with progressive myopia and routine optical biometry collected during 2015 –2026 was analysed. The Relative Asymmetry Index was defined per interval as the absolute interocular axial length difference normalised to the larger eye’s change; its patient-level median defined the Cumulative Relative Asymmetry Index (CRAI). An annualised Absolute Asymmetry Index (AAI) was introduced as a complementary rate metric.

**Results:** Median CRAI was 22.7 % [interquartile range 12.8–38.8]; median AAI 0.073 mm/year. AAI exceeded the patient-specific minimal detectable change (95 % confidence) in 18 % of patients (10–52 % across noise-estimate uncertainty); 13.9 % showed pronounced asymmetry (CRAI > 50 %; AAI 0.218 mm/year). CRAI rose with age (Spearman ρ = +0.28; partial ρ = +0.25 after adjustment for net axial length change rate, both p < 0.001), and was independent of the overall rate after age adjustment (partial ρ = −0.08, p = 0.20), consistent with scale invariance. Net axial length change rate decreased approximately three-fold across age strata (ρ = −0.23, p < 0.001).

**Conclusion:** CRAI and AAI provide complementary approximately scale-invariant and absolute metrics of interocular asymmetry. Interocular asymmetry appears common in this cohort and shows a marked age-related rise independent of overall progression rate.

## 1. Introduction

Myopia is among the most common ocular conditions in children worldwide, with prevalence estimated at approximately 30 % globally and exceeding 80 % in many East Asian populations.[1,2] Progressive axial elongation underlying childhood myopia is associated with substantial lifetime risk of vision-threatening complications including myopic maculopathy, retinal detachment, glaucoma, and high-myopia–associated optic neuropathy.[3,4] Slowing axial elongation has therefore become a primary objective of paediatric myopia management, supported by an expanding evidence base for pharmacological and optical interventions.[5–7]

Axial length (AL), measured by optical biometry, is widely accepted as the gold-standard objective marker of myopia progression in children.[8,9] Compared with cycloplegic refraction, AL is independent of accommodative tone and exhibits superior measurement reproducibility, making it particularly suitable for longitudinal monitoring.[10] The conventional approach to AL-based monitoring in clinical practice summarises progression by a single rate of axial elongation — typically the annualised change in mean binocular AL — benchmarked against age-stratified normative growth curves;[8] research practice variously uses per-eye or right-eye outcomes.[5,10]

This bilaterally averaged approach implicitly assumes that the two eyes progress in concer t. In practice, fellow eyes need not change at identical rates, and the discrepancy between them — the interocular asymmetry of axial elongation — has received little quantitative attention in paediatric populations. In adults, interocular AL asymmetry predicts accelerated progression and high-myopia outcomes,[11] yet its development during childhood remains poorly characterised. Standardised, scale-invariant metrics of asymmetric progression are not currently available in the paediatric myopia literature.

To address this gap, a relative asymmetry framework was developed and applied to a retrospective cohort of 267 children with progressive myopia followed for up to a decade. The Relative Asymmetry Index (RAI) is defined at the interval level, aggregated as the patient-level Cumulative Relative Asymmetry Index (CRAI), and complemented by the annualised Absolute Asymmetry Index (AAI). The aims were threefold: (i) to define and characterise the framework as a paired metric system for interocular asymmetry; (ii) to characterise CRAI prevalence across clinically meaningful categories; and (iii) to examine the age-related dynamics of CRAI and AAI relative to overall progression rate.

## 2. Methods

### 2.1. Study design and setting

This was a retrospective cohort study of anonymised longitudinal clinical records of paediatric patients with progressive myopia, collected during routine ophthalmological care at Eurostyle Clinic (Barnaul, Russian Federation) between 2015 and 2026. Clinical data were entered by treating clinicians in the course of routine practice using a structured digital ophthalmology platform (Myopia Tracker). No additional interventions or patient contacts were performed for the purposes of this study. The study was reported in accordance with the Strengthening the Reporting of Observational Studies in Epidemiology (STROBE) statement.[12]

### 2.2. Participants

Eligibility required at least two visits with optical biometry of both eyes during the study period at ages 5–16 years, and a diagnosis of progressive myopia. Patients with cataract, retinal disease, congenital ocular anomalies, prior intraocular or refractive surgery, or any condition precluding reliable biometry were excluded, as were patients with missing demographic data. From 283 initially identified patients, 16 were excluded (no biometry visit at age 5–16: 3; missing sex: 6; interval-level filtering failure: 7), yielding a final cohort of 267 patients contributing 913 valid follow-up intervals (Online Resource 1, Supplementary Figure S1). Patients received various myopia control therapies (including low-dose atropine, orthokeratology, defocus-incorporating soft contact lenses, and defocus-incorporating spectacle lenses) prescribed by their clinicians per routine practice; therapy assignment was non-randomised and heterogeneous; therapy-stratified description in Supplementary Table S6; detailed modelling reserved for a subsequent investigation.

### 2.3. Data sources and measurements

Axial length (AL) was measured at every visit by non-contact optical biometry (partial coherence interferometry or swept-source optical coherence biometry). All measurements of an individual patient were obtained on a single device throughout follow-up. Current optical biometers show within-session SD ≈ 0.014 mm for IOLMaster 700,[13] and same-day SD ≈ 0.04–0.06 mm across major instruments.[14,15] Diurnal AL variation (∼ 0.045 mm)[16] adds to inter-visit single-eye variability but is largely shared between fellow eyes. Keratometry was measured at every visit by autorefractor-keratometer. The analytical dataset comprised, per visit, a sequential numeric identifier independent of any platform key, age, AL of both eyes, and average keratometry; re-identification is not technically feasible.

### 2.4. Variables and intervals

For each pair of consecutive visits, an *interval* was defined by its duration in days, the AL changes of each eye (ΔAL_OD for the right eye, ΔAL_OS for the left eye) and their absolute difference. Baseline AL was the mean of OD and OS values at the first visit. The annualised net AL change rate was defined per interval as the signed change in AL at the eye with the larger absolute change, divided by interval duration in years; the patient-level rate was defined as the median across intervals. The signed formulation preserves the distinction between net axial elongation (positive, reflecting scleral growth) and net axial shortening (negative, reflecting choroidal thickening response to therapy exceeding scleral elongation); this sign carries opposite clinical significance in paediatric myopia management.

### 2.5. Asymmetry metrics

For each valid interval, the Relative Asymmetry Index (RAI) was defined to quantify the relative discordance between longitudinal AL changes of the two eyes:

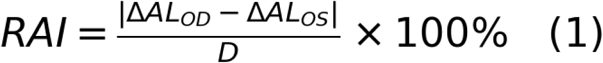

where *D* is defined piecewise to keep RAI bounded in [0 %, 100 %]: *D* = max(ΔAL_OD, ΔAL_OS) when both changes are non-negative (the typical progressive-myopia case); *D* = |min(ΔAL_OD, ΔAL_OS)| when both are negative (transient AL decrease, e.g. due to choroidal thickening); and *D* = |ΔAL_OD| + |ΔAL_OS| when the signs are opposite, yielding RAI = 100 % as a mathematical identity. RAI = 0 % corresponds to perfect concordance; RAI = 100 % to maximal discordance.

The patient-level **Cumulative Relative Asymmetry Index (CRAI)** was defined as the median of RAI across all valid intervals of a patient:

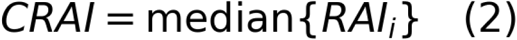

As a complementary metric, the annualised **Absolute Asymmetry Index (AAI)** was defined at the interval level as the absolute interocular difference in AL change per year, aggregated at the patient level by the median:

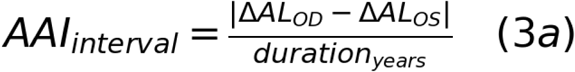

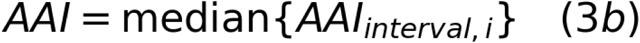

AAI is expressed in mm/year and directly comparable with the annualised net AL change rate. The two metrics capture complementary aspects: CRAI describes the relative pattern (approximately scale-invariant to overall axial change) and ranges from 0 % (concordant progression) to 100 % (maximal discordance), while AAI describes the absolute rate of asymmetry accumulation. Robustness is examined in Section 3.6 and Supplementary Methods.

### 2.6. Filtering criteria

Two interval-level filters were applied to ensure retained intervals reflect biologically meaningful axial change rather than measurement noise. An interval was retained only if max(|ΔAL_OD|, |ΔAL_OS|) ≥ 0.05 mm (approximately the standard error of inter-visit AL measurements[14,15]) and its duration was ≥ 90 days. The 0.05 mm criterion balances data preservation and noise rejection, and aligns with the threshold for clinically significant axial change adopted in recent multicentre paediatric myopia literature;[17] sensitivity analyses with thresholds 0.07 mm and 0.10 mm are reported in Section 3.6 and Supplementary Table S1. Of 1,128 raw visit pairs (consecutive visits at ages 5–16 within the eligible cohort), application of these filters yielded 913 valid intervals contributed by 267 patients (median 3 intervals per patient, IQR 1–5; Supplementary Figure S1). The 0.05 mm threshold operates as a single-eye, interval-level filter, distinct from the patient-level MDC₉₅ used to benchmark annualised AAI (Section 2.7).

### 2.7. Statistical analysis

Continuous variables are summarised as median [IQR] given the skewed distributions. Group comparisons used the Mann–Whitney U or Kruskal–Wallis test; bivariate associations used Spearman rank correlation. To distinguish direct from indirect effects in the presence of correlated predictors (notably age, baseline AL, and net AL change rate), partial Spearman correlations were computed via rank regression: the rank-transformed predictor and outcome were each linearly regressed on the rank-transformed control variable(s), and the Pearson correlation of the resulting residuals was used as the partial coefficient. Within- and between-patient components of interval-level associations were analysed by patient-mean centring. Prevalence estimates are reported with 95 % Wilson confidence intervals. To benchmark patient-level AAI against measurement uncertainty, the visit-level measurement-error SD of the interocular AL difference (σ_e_) was estimated directly from the cohort as the square root of the negative lag-1 autocovariance of consecutive signed interval-level differences, which share a visit (Supplementary Methods); the noise floor and a patient-specific MDC₉₅[19,20] were then obtained by simulation from the null distribution of AAI under each patient’s observed interval structure. Robustness of principal findings (noise threshold, aggregator, denominator definition, duration filter, category cut-offs) was assessed by sensitivity analyses (Section 3.6, Supplementary Methods and Tables S1–S3). A total of 22 formal hypothesis tests are reported; principal age-related findings (both p < 10⁻⁴) survive Bonferroni correction at any reasonable N, while the AAI-by-age Kruskal–Wallis result (p = 0.01) is reported as supportive.

### 2.8. Ethics statement

The Local Ethics Committee of Eurostyle Clinic granted a waiver of formal ethics review (No. LEC-26-07/3, 15 July 2026); individual informed consent was not required (see Declarations).

## 3. Results

### 3.1. Cohort characteristics

The cohort comprised 267 paediatric patients with progressive myopia (150 girls [56.2 %], 117 boys [43.8 %]), contributing 913 valid intervals (median 3 per patient). Median age at first visit was 8.1 years [IQR 6.7–9.8] and median follow-up 1.71 years [0.71–3.12]. Median mean baseline AL was 23.77 mm [23.06–24.57]; baseline interocular AL difference was small (median 0.09 mm [0.04–0.18]), with axial anisometropia (> 0.35 mm, ≈ 1 D) in 8.2 % at baseline. Median annualised net AL change rate was 0.28 mm/year [0.20–0.40], confirming an actively progressing cohort (Table 1). The minimal baseline interocular AL asymmetry contrasts with the substantial asymmetry in net AL change rates examined below, supporting an interpretation of progression asymmetry as a dynamic phenomenon distinct from baseline anatomical asymmetry.

**Table 1.** Cohort characteristics (n = 267). Demographics, follow-up parameters, baseline biometry, and overall annualised net AL change rate of the analytical cohort. Continuous variables are summarised as median [interquartile range].

| Characteristic | Value |
| --- | --- |
| *Demographics* |  |
| Female, n (%) | 150 (56.2) |
| Male, n (%) | 117 (43.8) |
| Age at first visit, years, median [IQR] | 8.1 [6.7–9.8] |
| Age at last visit, years, median [IQR] | 10.6 [9.1–11.9] |
| *Follow-up* |  |
| Follow-up duration, years, median [IQR] | 1.71 [0.71–3.12] |
| Total visits per patient, median [IQR; range] | 4 [3–7; 2–19] |
| Valid analysis intervals per patient, median [IQR; range] | 3 [1–5; 1–13] |
| Patients with a single valid interval, n (%) | 71 (26.6) |
| *Baseline biometry* |  |
| Mean baseline axial length, mm, median [IQR] | 23.77 [23.06–24.57] |
| Range of mean baseline axial length, mm | 21.44–26.49 |
| Baseline interocular AL difference, mm, median [IQR] | 0.09 [0.04–0.18] |
| Axial anisometropia (interocular AL difference > 0.35 mm, ≈ 1 D), n (%) | 22 (8.2) |
| Keratometry available, n (%) | 232 (86.9) |
| Average keratometry, D, median [IQR] | 43.25 [42.50–44.25] |
| *Progression* |  |
| Annualised net AL change rate, mm/year, median [IQR] | 0.28 [0.20–0.40] |

IQR — interquartile range; AL — axial length; D — dioptres.

### 3.2. Prevalence of interocular asymmetry of myopia progression

Across the 913 valid intervals, median RAI per interval was 21.4 % [IQR 10.0 –42.9]. At the patient level, median CRAI was 22.7 % [12.8–38.8] and median AAI 0.073 mm/year [0.038–0.114] (Fig. 1).

**Fig. 1.**
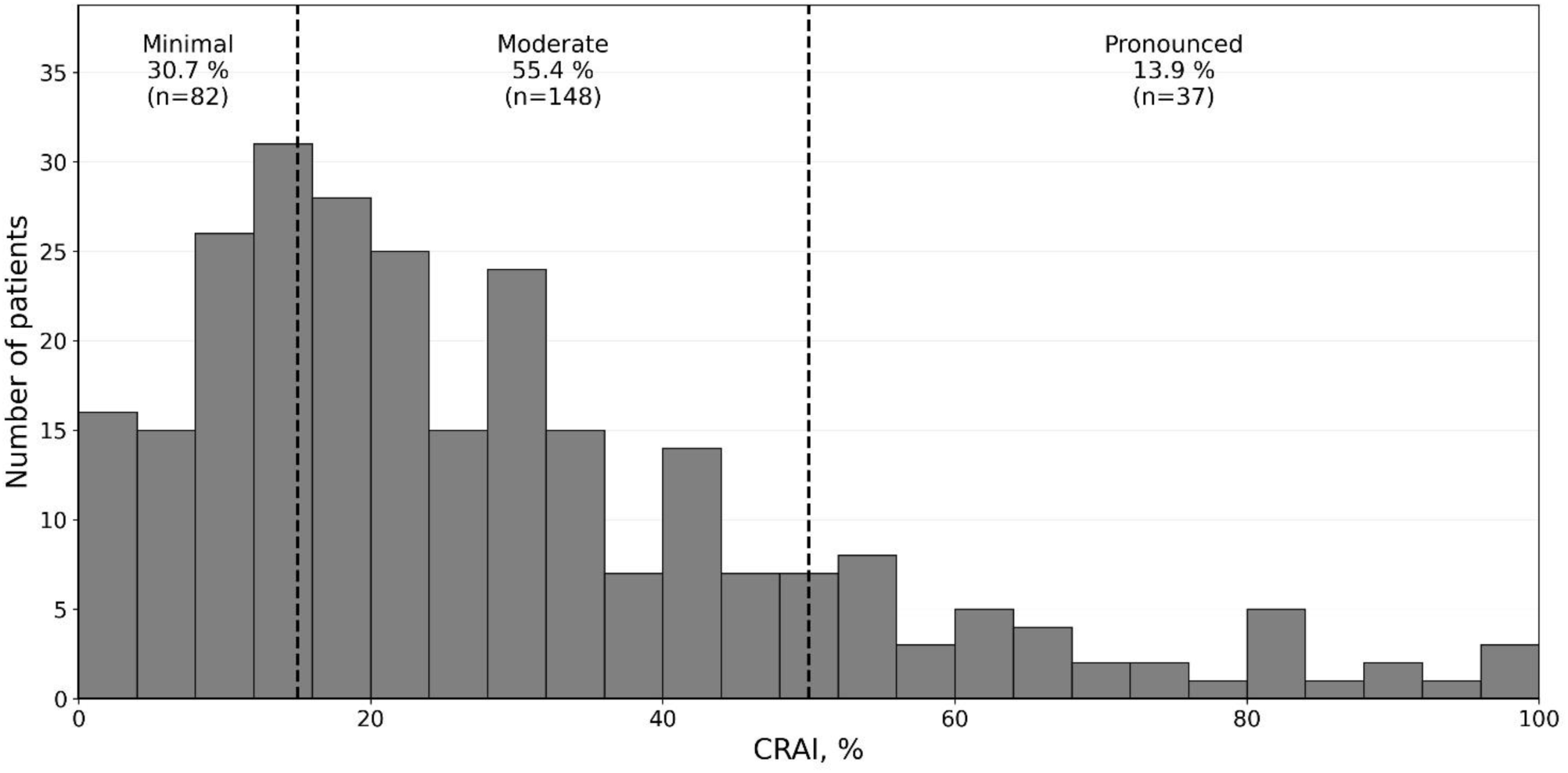
Distribution of the Cumulative Relative Asymmetry Index (CRAI) and prevalence of asymmetry categories in the analytical cohort (n = 267). Histogram of patient-level CRAI values (in %), with category cut-offs at 15 % and 50 % shown as vertical dashed lines and category fractions labelled. The distribution is right-skewed (skewness 1.23), with approximately 70 % of patients exceeding the 15 % threshold for non-minimal asymmetry, and 13.9 % (95 % Wilson CI 10.2–18.5) exceeding the 50 % threshold for pronounced asymmetry.

To facilitate clinical interpretation, three asymmetry categories were defined with empirically motivated thresholds: minimal asymmetry (CRAI ≤ 15 %), moderate (> 15–50 %), and pronounced (>50 %). The 15 % threshold is a statistical noise filter: 88 % of patients in this stratum have AAI below the patient-level measurement noise floor. The 50 % threshold has a direct biological interpretation: at RAI ≥ 50 %, the slower-progressing eye accumulates axial length at no more than half the rate of the faster-progressing eye over the interval — a clinically substantial discordance. Minimal asymmetry was observed in 30.7 % (n = 82; 95 % Wilson CI 25.5–36.5), moderate in 55.4 % (n = 148; 49.4–61.3), and pronounced in 13.9 % (n = 37; 10.2–18.5) — Table 2.

**Table 2.** Prevalence of interocular asymmetry of myopia progression by CRAI category in the analytical cohort (n = 267). Categories are defined by patient-level Cumulative Relative Asymmetry Index (CRAI). The pronounced-asymmetry stratum corresponds to a clinically substantial discordance where the slower-progressing eye accumulates axial length at no more than half the rate of the faster-progressing eye over the median interval, with median AAI approximately 3.8-fold above the patient-level noise floor (∼0.057 mm/year). Independently, 18 % of the cohort has individual AAI exceeding its patient-specific Minimal Detectable Change at 95 % confidence (MDC₉₅; median 0.138 mm/year, range 0.020 –0.336 across patients, reflecting differences in the number and duration of contributing intervals); these two complementary criteria — clinical severity (CRAI > 50 %) and statistical detectability (AAI > MDC₉₅) — provide convergent support for the reality of meaningful asymmetry. 95 % Wilson confidence intervals are reported for prevalence estimates.

| Category | CRAI range | n | % (95 % Wilson CI) | Median AAI, mm/year | % below noise floor (0.057) | % above patient specific MDC<sub>95</sub> |
| --- | --- | --- | --- | --- | --- | --- |
| Minimal asymmetry | ≤ 15 % | 82 | 30.7 (25.5–36.5) | 0.026 | **88** | 1 |
| Moderate asymmetry | > 15–50 % | 148 | 55.4 (49.4–61.3) | 0.085 | 22 | 15 |
| Pronounced asymmetry | > 50 % | 37 | 13.9 (10.2–18.5) | 0.218 | **0** | 70 |
| **Total** |  | **267** | **100.0** | 0.073 | 39 | 18 |

### 3.3. Characterisation of the asymmetry metric

Four properties of the asymmetry metric were assessed (Supplementary Methods and Tables S4–S5). First, scale invariance: by construction, CRAI quantifies the asymmetric fraction of axial elongation per interval, normalised to the change of the faster-progressing eye. Consistent with this interpretation, partial Spearman correlation between CRAI and the annualised net AL change rate, after adjustment for age, was not significantly different from zero (partial ρ = −0.08, p = 0.20), consistent with approximate scale invariance of CRAI. This test is not tautological despite CRAI and the net rate sharing the denominator term: simulations calibrated to the observed CRAI level yield partial ρ = +0.08 when the asymmetric component is proportional to total elongation and −0.27 when it is fixed in absolute terms (Supplementary Methods), so the observed value discriminates between these regimes. By contrast, AAI — the absolute interocular difference, not self-normalised to within-interval amplitude — showed a strong positive association with net AL change rate (partial ρ = +0.35, p < 0.001 after age adjustment), as expected for an absolute rate metric.

CRAI and AAI thus capture complementary information: the relative pattern (independent of overall amplitude) and the absolute magnitude of interocular asymmetry. Second, within-patient variability: ∼42 % of interval-level RAI variance was between-patient (descriptive); the patient-level REML ICC was 0.18[21] (Supplementary Methods). The observed lag-1 autocorrelation of the signed interocular difference was −0.18 (p < 10⁻⁵), incompatible with measurement noise alone, which predicts −0.5 because consecutive intervals share a visit (Supplementary Methods). Its autocovariance across all raw consecutive visit pairs gave σ_e_ = 0.030 mm (cluster-bootstrap 95 % CI 0.015–0.044; lag-2 autocorrelation +0.04). CRAI summarises central tendency at the patient level. Third, residual dependence on interval length: after the 90-day duration filter, RAI retained a weak negative association with interval duration (ρ = −0.13, p < 0.001). Because intervals are clustered within patients, this association was decomposed by patient-mean centring: it was present both within patients (r = −0.10, p = 0.004) and between patients (ρ = −0.15, p = 0.02), and remained significant under a cluster bootstrap (p < 0.001). Impact under a more conservative 180 -day threshold is examined in Section 3.6. Fourth, robustness to boundary intervals: excluding RAI = 100 % intervals preserved the age-related rise (ρ = +0.243, p < 10⁻⁴; Supplementary Methods).

### 3.4. Age-related dynamics: CRAI and AAI

CRAI correlated positively with age at first visit (Spearman ρ = +0.28, p < 0.001). Because the annualised net AL change rate is itself negatively correlated with age (ρ = −0.23, p < 0.001), consistent with the well-described deceleration of axial elongation through later childhood,[8,18] partial correlations were used to disentangle the two effects. After adjustment for net AL change rate, CRAI retained a significant positive association with age (partial ρ = +0.25, p < 0.001; +0.22, p < 0.001 with additional adjustment for follow-up duration), and AAI likewise showed a positive partial association (partial ρ = +0.32, p < 0.001). Because CRAI aggregates intervals spanning the whole follow-up while age was defined at entry, the association was verified at the interval level, where no such mismatch arises: RAI rose with age at the start of each interval both across intervals (ρ = +0.15, p < 0.001) and within patients (r = +0.10, p = 0.006; cluster bootstrap p < 0.001).

Across age strata (Table 3, Fig. 2), CRAI rose from 19.2 % at 5–8 years to 33.3 % at 10–12 years, with a slight decrease to 32.0 % at 12–16 years; AAI rose from 0.060 to 0.106 mm/year, while the annualised net AL change rate decreased approximately three-fold (0.29 → 0.09 mm/year). Kruskal–Wallis tests confirmed significant age differences for CRAI (H = 21.1, p < 0.001), AAI (H = 11.3, p = 0.01), and net AL change rate (H = 21.5, p < 0.001).

**Table 3.** Age-related dynamics of asymmetry metrics and net AL change rate (n = 267). Patient-level CRAI, annualised AAI, and the overall annualised net AL change rate are presented across four age strata defined by age at first visit. Kruskal–Wallis tests assess differences across strata.

| Age stratum, years | n | CRAI, %, median [IQR] | AAI, mm/year, median [IQR] | Annualised net AL change rate, mm/year, median [IQR] |
| --- | --- | --- | --- | --- |
| 5–8 | 128 | 19.2 [11.1–29.9] | 0.060 [0.030–0.098] | 0.291 [0.235–0.435] |
| 8–10 | 84 | 28.2 [14.2–41.1] | 0.088 [0.042–0.117] | 0.276 [0.202–0.382] |
| 10–12 | 43 | 33.3 [21.9–51.8] | 0.087 [0.059–0.114] | 0.223 [0.150–0.302] |
| 12–16 | 12 | 32.0 [12.8–74.2] | 0.106 [0.045–0.157] | 0.088 [–0.138–0.311] |
| *Kruskal–Wallis p value* | | $< 0.001$ | 0.010 | $< 0.001$ |

CRAI — Cumulative Relative Asymmetry Index; AAI — Absolute Asymmetry Index (annualised); AL — axial length. Age strata are left-closed (e.g. 8–10 denotes $8.0 \leq \text{age} < 10.0$ ); no patient fell exactly on a stratum boundary. Bootstrap 95 % confidence intervals (10,000 resamples) for stratum medians of CRAI: 16.7–21.8 % (5–8 yr), 18.2–31.5 % (8–10 yr), 25.8–42.4 % (10–12 yr), 12.7–76.9 % (12–16 yr); for stratum medians of AAI: 0.048–0.068, 0.070–0.096, 0.075–0.099, 0.040–0.170 mm/year respectively.

**Fig. 2.**
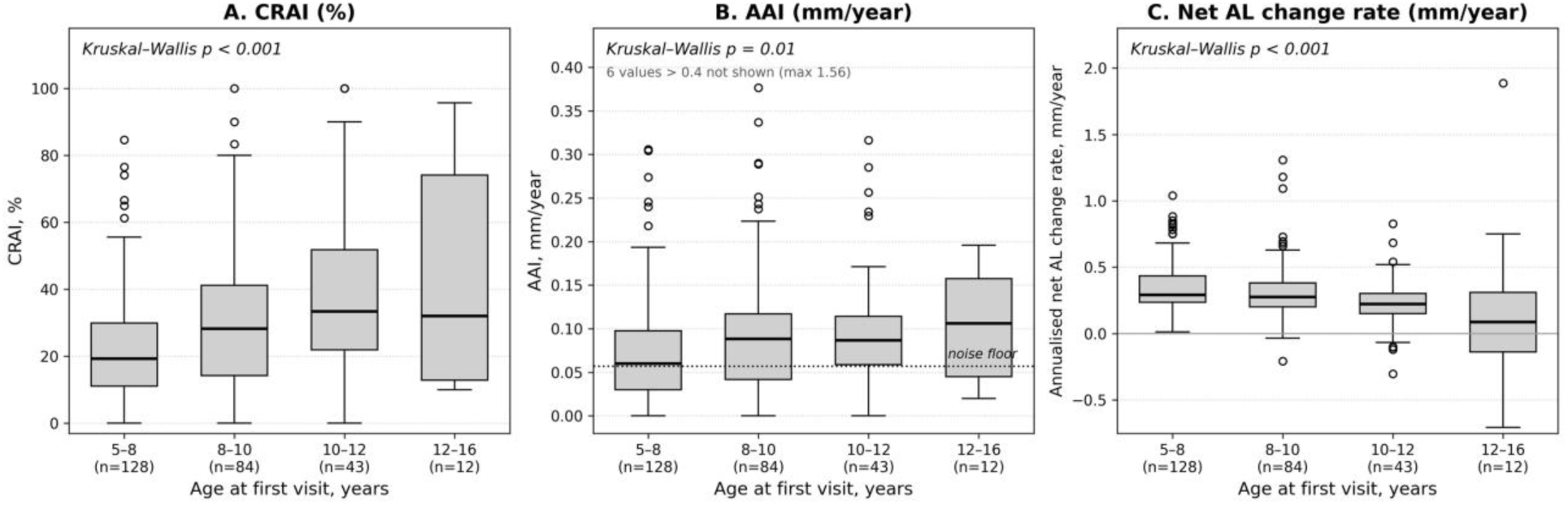
Age-related dynamics of asymmetry metrics in contrast to the overall net AL change rate (n = 267). Patient-level CRAI (panel A), annualised AAI (panel B), and the overall annualised net AL change rate (panel C) are shown across four age strata (5 –8 years, 8–10 years, 10–12 years, 12–16 years; n = 128, 84, 43, 12 respectively). Box-and-whisker plots show median, interquartile range, and 1.5 × IQR whiskers. In panel B the ordinate is truncated at 0.4 mm/year for legibility; six patients with higher values (maximum 1.56 mm/year) are not displayed, and the dotted line marks the patient-level noise floor (0.057 mm/year). CRAI rises from 19.2 % to a peak of 33.3 % at 10–12 years with a slight decrease to 32.0 % at 12–16 years, AAI rises (from 0.060 to 0.106 mm/year), while the net AL change rate decreases approximately three-fold. Kruskal–Wallis tests confirm significant differences across age strata for all three metrics (p ≤ 0.01).

The age-related rise of CRAI is not an artefact of decreasing net AL change rate: AAI also rises with age, and the proportion of patients exceeding their patient-specific MDC₉₅ does not decline across age strata (14.8 %, 20.2 %, 20.9 %, 33.3 %). The proportion of patients below the patient-level noise floor decreases from 48 % at age 5–8 to 23–33 % in the two oldest strata, indicating that more patients show statistically detectable asymmetry in older strata, not fewer.

### 3.5. Associations with sex, baseline AL, and keratometry

No significant sex differences were observed for CRAI (girls 21.8 % [IQR 12.5 –34.5] vs boys 25.0 % [14.3–41.9], p = 0.22) or AAI (p = 0.62). CRAI showed a modest positive bivariate correlation with baseline AL (ρ = +0.20, p = 0.001), but baseline AL was strongly correlated with age (ρ = +0.54) and, after age adjustment, the association became non-significant (partial ρ = +0.06, p = 0.31); baseline AL does not independently predict asymmetry beyond age. Average keratometry showed no association with either metric (n = 232; CRAI ρ = +0.00, p = 0.97; AAI ρ = −0.02, p = 0.75). Age at first visit was thus the principal demographic-biometric determinant of interocular asymmetry.

### 3.6. Sensitivity analyses

The robustness of the principal findings to methodological choices was assessed. Varying the noise threshold (0.05, 0.07, 0.10 mm) yielded patient-level CRAI values strongly correlated with the principal specification (ρ ≥ 0.90), with category assignment preserved in 87 –100 % of patients. Alternative aggregators (arithmetic mean, 10 % trimmed mean) showed strong agreement with the median (ρ ≥ 0.93; ≥ 88 % concordance). Alternative RAI denominators (sum, mean of absolute AL changes) preserved patient ranking essentially unchanged (ρ ≥ 0.99). A more conservative 180-day duration filter reduced the cohort to 206 patients (481 intervals); median CRAI shifted modestly to 20.2 %, and pronounced-asymmetry prevalence remained essentially unchanged (14.6 %). The age-related rise was attenuated and became marginally non-significant in this smaller subset after adjustment (partial ρ = +0.13, p = 0.07), likely driven by reduced power. Detailed results in Supplementary Tables S1–S3.

## 4. Discussion

To the author’s knowledge based on PubMed searches in July 2026, this is the first dedicated metric framework for asymmetric axial elongation progression in paediatric myopia, distinct from cross-sectional reports of static anisometropia. In a cohort of 267 children with progressive myopia followed for up to a decade of routine clinical care (median follow-up 1.71 years), 18 % exhibited statistically detectable interocular asymmetry at the individual level (AAI exceeding their patient-specific MDC₉₅; median 0.138 mm/year), and 13.9 % demonstrated pronounced asymmetry — defined by CRAI > 50 %, where the slower-progressing eye accumulates axial length at no more than half the rate of the faster-progressing eye. Beyond these two groups, the majority displayed a non-minimal asymmetric component (CRAI > 15 %; ∼70 % of patients), though this relative criterion is less stringent than statistical detectability. In the pronounced-asymmetry stratum, median AAI was approximately 3.8-fold above the patient-level noise floor, no patients fell below it, and 70 % exceeded MDC₉₅ — combining clinical severity (CRAI > 50 %) with high statistical detectability.

Together, these two complementary perspectives — clinical severity at the patient level (CRAI > 50 %) and statistical detectability at the individual level (AAI > MDC₉₅) — provide convergent evidence that meaningful interocular asymmetry of axial elongation is a real, measurable feature of paediatric myopia progression rather than measurement variation. While interocular AL asymmetry is recognised as a risk marker for accelerated myopia progression in adults,[11] its prevalence and dynamic behaviour during childhood have not previously been quantified using a self-normalised metric.

The age-related dynamics observed in this cohort can be interpreted within a two -component framework of net axial change. The net AL change rate decreased approximately three-fold from early childhood to adolescence, reflecting both the well-described developmental deceleration of scleral elongation[8,18] and, in older strata, choroidal thickening response to myopia control interventions — a response with bilateral and potentially asymmetric components — that can produce net AL shortening exceeding scleral elongation over a follow-up interval. Against this background, both CRAI and AAI increased with age, even after adjustment for net AL change rate.

This pattern suggests two distinct contributions to axial elongation: a common bilateral growth component, plausibly reflecting bilateral emmetropisation pressure, systemic hormonal regulation, and shared environmental drivers, which fades with maturation; and a local asymmetric component, plausibly reflecting eye-specific differences in accommodative load, near-work posture, lateralised choroidal response, or interocular vergence imbalance, which appears to remain stable or to intensify modestly across childhood. This common-versus-asymmetric framework is therefore supported by three independent age-related trajectories — declining net AL change rate, rising CRAI, and rising AAI — that are mutually consistent rather than tautologically linked. Age-related differences were observed cross-sectionally across age strata; within-subject longitudinal verification with denser sampling is reserved for future work and acknowledged in the limitations.

The modest age-related intensification of the asymmetric component is consistent with the developmental trajectory of paediatric visual behaviour: schoolwork, reading, and digital device use increase markedly with age, often in laterally asymmetric postures (head tilt, dominant-hand device-holding, monocular near-work) that produce asymmetric accommodative demands. Because accommodative lag drives hyperopic defocus and axial elongation in myopic children, sustained interocular differences in lag could translate into persistent excess elongation in the eye with greater lag. However, not all such children develop measurable asymmetry, implicating genetic susceptibility as a modifier — a hypothesis for future genetic-epidemiological study. Ocular dominance was not assessed in the present retrospective cohort and is planned for prospective characterisation alongside leading-eye dynamics.

Under this framework, CRAI can be read as an estimate of the proportion of total axial elongation attributable to the asymmetric component: as the common component declines, the relative weight of the asymmetric component within total observed axial change grows, producing the observed age-related rise of CRAI in older children even when its absolute magnitude rises only modestly. Seasonal modulation may contribute to within-patient variability; leading-eye persistence and switching dynamics are reserved for a follow-up study.

Notably, this dynamic asymmetry emerges against minimal baseline axial anisometropia (median AL difference 0.09 mm; 95th percentile 0.41 mm, near the ≈ 1 D equivalent). Whether longitudinal accumulation produces clinically meaningful anisometropia is reserved for future investigation.

Clinical implications are threefold. First, monitoring should track each eye separately rather than rely on bilaterally averaged AL, since one in seven children shows pronounced asymmetry. Second, the pronounced-asymmetry stratum (CRAI > 50 %; AAI clearly above the noise floor) is clinically distinguishable and may warrant particular attention. Third, clinicians should expect increasing asymmetry in older children even as the net AL change rate decreases — an evolving balance between two growth components, not worsening disease.

Whether established myopia control therapies[5,6] act differentially on the common and asymmetric components is an open question that the present descriptive framework cannot address but is well positioned to motivate.

A broader methodological implication concerns global myopia research practice. Existing landmark myopia control trials[5–7] have characterised treatment efficacy through outcomes — whether single-eye analysis under monocular treatment allocation,[5] right-eye analysis,[7] or per-eye reporting under bilateral treatment[6] — that cannot express interocular asymmetry of progression.

Even where asymmetry averages toward zero across follow-up — for instance through switching of the leading eye between sides — the underlying dynamic process, plausibly reflecting asymmetric accommodative load, lateralised choroidal response, or interocular vergence imbalance, is discarded by bilaterally aggregated outcomes rather than shown to be absent. Whether asymmetry-aware analysis of existing trial datasets would revise or refine efficacy estimates, and whether asymmetry-targeted intervention improves outcomes, are open empirical questions that the metrics proposed here make tractable.

The retrospective observational design cannot establish causality. Category thresholds (15 %, 50 %) were partially data-informed rather than pre-specified. The cohort originates from a single regional population; external validation in other paediatric populations is required before prevalence figures are generalised. Intra-patient measurements were obtained on a single device throughout follow-up, but exact device identification was not consistently available. Stratum-specific noise-floor relationships are detailed in Table 2 and Supplementary Methods. AAI and its MDC₉₅ characterise the rate at which interocular asymmetry is generated and are, by construction, insensitive to slow accumulation of a persistent same-sign difference: a patient may accumulate a clinically meaningful interocular difference through per-interval increments that individually fall below detectability. The cohort necessarily reflects patients engaged with structured digital monitoring, introducing potential selection bias. The patient-level reliability of CRAI as a stable individual trait is poor by conventional criteria[21] (REML ICC = 0.18), consistent with genuine within-patient dynamics of asymmetry rather than a fixed trait; cohort-level prevalence and age-related findings remain robust under sensitivity analysis restricted to patients with ≥ 2 valid intervals (Supplementary Methods). The 12 –16 year stratum is small (n = 12), yielding a wide bootstrap 95 % CI for median CRAI (12.7 –76.9 %); cohort-level findings remain robust. A post-hoc analysis (Supplementary Table S6) showed pronounced asymmetry distributed across all therapy subgroups, including 7.4 % of 122 patients with no active therapy (χ² p = 0.11).

Interocular asymmetry of axial elongation appears to be a common and dynamic feature of paediatric myopia progression. CRAI and AAI together provide approximately scale-invariant and absolute metrics that complement net AL change rate. The age-related rise of CRAI alongside a decreasing net AL change rate is consistent with a two-component model. Future work should address prospective dynamics, treatment response, and external validation.

## Declarations

### Author Contributions

The author is the sole author and meets the ICMJE criteria for authorship: substantial contribution to conception and design, acquisition, analysis, and interpretation of data; drafting and critical revision of the manuscript; final approval of the version to be published; and accountability for all aspects of the work in ensuring that questions related to the accuracy or integrity of any part of the work are appropriately investigated and resolved.

### Data availability

The anonymised patient-level dataset (n = 267), the Python reference implementation of the asymmetry metrics, and a complete analysis pipeline reproducing the principal numerical findings are deposited on Zenodo (DOI: 10.5281/zenodo.20319399) under restricted access. Editors, peer reviewers, and qualified researchers with a defined methodological purpose may request access through the repository’s access-request mechanism, subject to the stated conditions. Running the deposited pipeline against the deposited dataset reproduces all principal numerical findings reported in the manuscript. Interval-level raw axial length measurements are not shared in order to minimise the residual risk of patient re-identification, but are available from the corresponding author upon reasonable request from researchers with a defined methodological purpose.

### Ethics approval

The study was reviewed by the Local Ethics Committee of Eurostyle Clinic (Barnaul, Russian Federation), the institution where data collection occurred, which granted a waiver of formal ethics review (waiver No. LEC-26-07/3 dated 15 July 2026) given the fully retrospective nature of the analysis on anonymised clinical records. The waiver confirms compliance with the principles of the Declaration of Helsinki and applicable national legislation on personal data protection and medical confidentiality. The committee determination letter is available upon editorial request.

### Consent to participate

This retrospective study used anonymised clinical records collected during routine care. The Local Ethics Committee of Eurostyle Clinic waived the requirement for individual informed consent given the retrospective design and the use of fully anonymised data (waiver No. LEC-26-07/3).

### Consent to publish

This article does not contain any individual person’s identifiable data, images, or videos; consent to publish was therefore not required.

### Use of artificial intelligence

Artificial intelligence assistance (Anthropic’s Claude Opus 4.6– 4.8, accessed via claude.ai during October 2025–July 2026) was used for language editing of the manuscript, verification of structural coherence and internal consistency across se ctions, and verification of statistical reasoning and computed results against the deposited code pipeline (see Data availability). All scientific content, study design, choice of statistical methods, data analyses, interpretations, and conclusions remain the responsibility of the author. The AI application did not contribute to data collection or analysis, and does not meet ICMJE criteria for authorship.

## Acknowledgments

None.

---

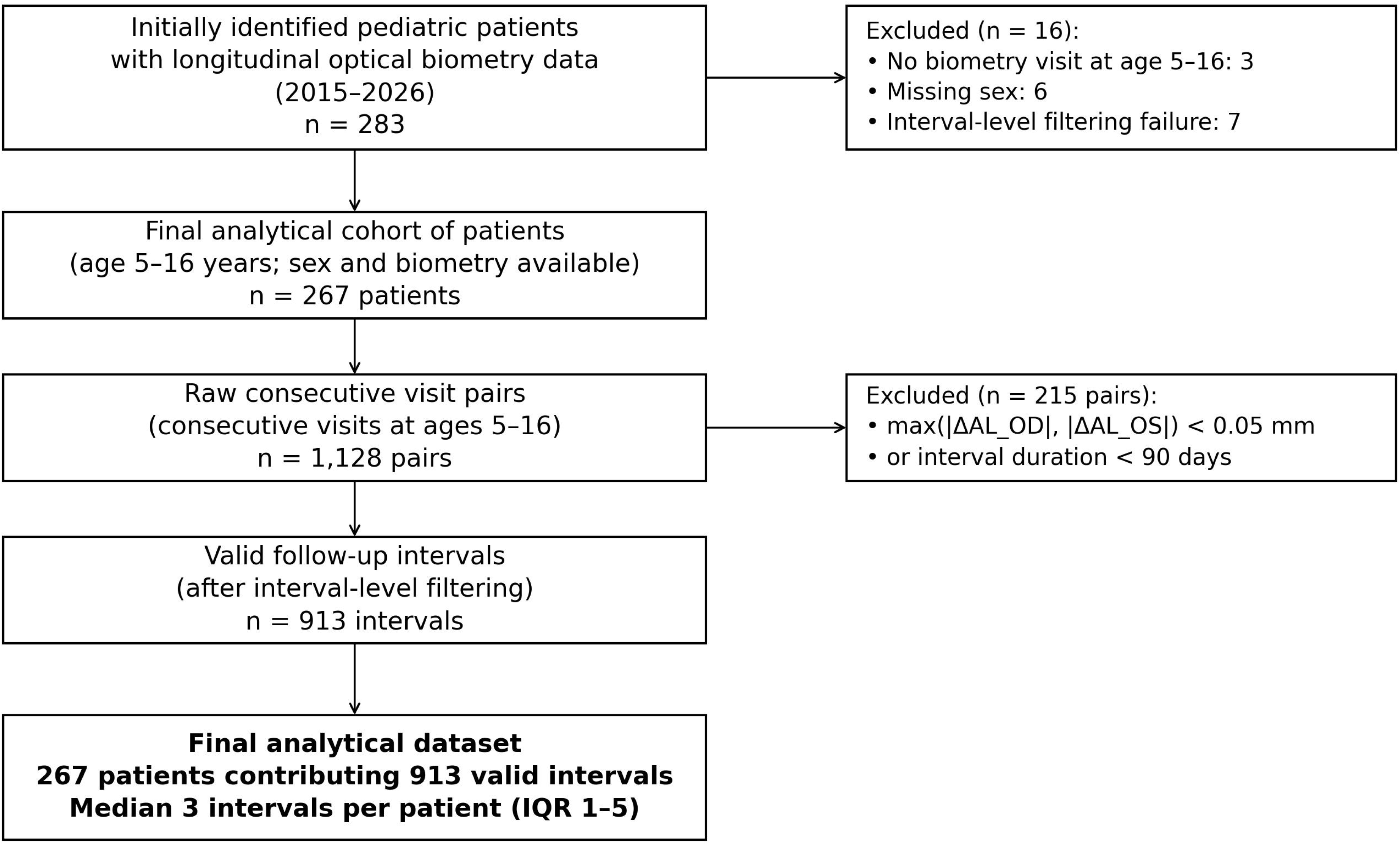

