## Supplementary Material for "Quantifying interocular asymmetry of axial elongation in childhood myopia: metric framework, prevalence, and age-related dynamics"

**Online Resource 1. Supplementary Material for: "Quantifying interocular asymmetry of axial elongation in childhood myopia: metric framework, prevalence, and age-related dynamics"**

Rodion Nesterenko, MD

Eurostyle Clinic, ul. Molodezhnaya 3b, Barnaul 656038, Russian Federation

**Contents**:

- Supplementary Methods
- Supplementary Tables S1–S7
- Supplementary Figure S1

### Supplementary materials

#### Supplementary Methods

*Derivation of the patient-level AAI noise floor and MDC₉₅.* The methodological framework follows established measurement-error theory in clinical research,^[19,20]^ adapted to longitudinal optical biometry. The inter-visit reproducibility of single AL measurements by optical biometry is conservatively estimated at SEM_AL ≈ 0.05 mm. This estimate is a **derived upper bound**, not a direct empirical measurement of between-day reproducibility (which is not well characterised in the paediatric myopia literature); it combines (i) within-session repeatability ranging from Sw ≈ 0.014 mm (IOLMaster 700 in children;^[13]^) to ≈ 0.022 mm (95 % LoA ±0.043 mm;^[14]^); (ii) same-day repeat intra-subject SDs of 0.04–0.06 mm across optical biometers;^[15]^ and (iii) diurnal physiological variation of axial length (~ 0.045 mm^[16]^). For an interval, the change ΔAL of one eye is the difference of two AL measurements, with SEM(ΔAL) = SEM_AL × √2 ≈ 0.071 mm, and the interocular difference |ΔAL_OD − ΔAL_OS| would, under fully independent measurement errors between fellow eyes, have SEM ≈ 0.10 mm per interval. The total SEM_AL ≈ 0.05 mm can be decomposed into an instrumental component (independent between fellow eyes, σ_inst ≈ 0.022 mm based on within-session repeatability of contemporary biometers[13,14]) and a diurnal-physiological component (bilaterally correlated within a single measurement session, σ_diur ≈ 0.045 mm[16]), satisfying σ_inst² + σ_diur² ≈ SEM_AL². Because the diurnal component is shared between fellow eyes at each visit and largely cancels in the interocular difference, the theoretical lower bound on the effective SEM of the interocular difference is 2σ_inst ≈ 0.044 mm per interval under fully correlated diurnal variation.

The effective SEM of the interocular difference was then estimated directly from the cohort rather than adopted as a working value (see Estimation of interocular measurement error below). The estimate, 0.043 mm per interval (σ_e_ = 0.030 mm per visit), coincides with the theoretical lower bound above; the previously adopted conservative value of 0.050 mm lies within its confidence interval.

Two features of the patient-level AAI make the conventional expression MDC₉₅ = 1.96 × SEM inapplicable to it. First, AAI is the median of the absolute interocular difference, whose null distribution is not centred at zero (for a signed difference distributed N(0, σ), the expected magnitude under pure noise is 0.674σ), whereas 1.96 × SEM is derived for a single signed measurement whose null is symmetric about zero. Second, σ/√n is the standard error of a mean and does not describe a median of half-normal variates. In addition, because each interval is annualised by its own duration, the null dispersion of AAI is interval-specific rather than common: with the estimated interval-level SEM of 0.043 mm and observed durations ranging from 0.25 to 5.89 years (median 0.50), the annualised null SD ranges from approximately 0.17 to 0.01 mm/year across intervals.

The noise floor and MDC₉₅ were therefore obtained directly from the null distribution of the statistic actually used, by Monte-Carlo simulation over the observed design (20,000 replicates). Independent measurement error was applied to axial length at each visit and separately to each eye, with a per-measurement SD of 0.0213 mm, so that the interocular difference has the estimated SD σ_e_ = 0.030 mm per visit and 0.043 mm per interval of change; this construction preserves the correlation between consecutive intervals, which share a visit. For each patient, simulated AAI was computed over that patient's own intervals, using their observed durations and their observed number of intervals.

This yields a cohort-level noise floor — the median of the null distribution — of 0.057 mm/year, and a patient-specific MDC₉₅, defined as the 95th percentile of each patient's own null distribution (median across patients 0.138 mm/year, interquartile range 0.112–0.171, range 0.020–0.336). The dispersion reflects the number and duration of a patient's contributing intervals: patients contributing a single short interval have an MDC₉₅ near 0.17 mm/year, whereas patients contributing eight or more intervals have one near 0.10 mm/year. Detectability was therefore assessed against each patient's own threshold rather than a common cohort value, so that patients with sparser follow-up are not credited with detectable asymmetry on the strength of a noisier estimate.

The noise floor is robust to these design differences (a fixed-σ approximation evaluated at the median duration and median interval count gives 0.057 mm/year), whereas the 95th percentile is not: the same approximation gives 0.127 mm/year against 0.166 mm/year for a single cohort-level threshold derived from the observed design, because the upper tail of the null is dominated by patients contributing few and short intervals. For reference, 1.96 × σ/√3 ≈ 0.100 mm/year would correspond to a false-positive rate of approximately 20 % rather than 5 %, and was not used.

*Sensitivity analysis design.* Sensitivity analyses were performed by re-running the principal analytical pipeline under alternative methodological choices, with the principal specification serving as the reference. For each variant, agreement with the principal specification was quantified by the Spearman correlation of patient-level CRAI values (ρ_CRAI) and by the percentage of patients assigned to the same asymmetry category under the principal cut-offs (≤ 15 %, 16–50 %, > 50 %).

*Comparison of cut-off systems.* The principal cut-offs were chosen empirically based on the relationship between CRAI and the annualised AAI (Supplementary Table S5). For comparison, two alternative cut-off systems are reported: a convention-based 10 % / 30 % scheme, and an intermediate 15 % / 45 % scheme.

*Therapy classification for post-hoc stratification.* For the post-hoc therapy-stratified description (Supplementary Table S6), each patient's primary therapy was defined as the category of the therapy episode of greatest duration during the analysed follow-up period. Ten source categories from clinical records (no_therapy; atropine_mono; atropine_plus_ortho; atropine_plus_soft; atropine_plus_defocus_glasses; ortho_mono; soft_lens_mono; defocus_glasses_mono; multi_optical; other_combination) were grouped into six analytical groups to balance specificity and statistical power: (1) no active therapy; (2) atropine monotherapy; (3) atropine plus non-orthokeratology optical (defocus-incorporating soft contact lenses or defocus-incorporating spectacle lenses); (4) orthokeratology (alone or combined with atropine); (5) optical monotherapy (non-orthokeratology); and (6) other or multi-optical combinations. Group 4 was specified as a distinct stratum because orthokeratology induces transient anterior-segment changes that could theoretically influence interocular asymmetry estimates derived from axial biometry, and a separate stratum allowed direct examination of this concern. Differences in CRAI category proportions across the six therapy groups were tested by Pearson's χ² test; differences in CRAI and AAI distributions between patients with and without orthokeratology as primary therapy were tested by the Mann–Whitney U test. The stratification is descriptive; the study was not designed to estimate therapy effects.

*Multiple-testing inventory.* The 22 formal hypothesis tests reported comprise: Spearman bivariate and partial correlations (n = 13; Sections 3.3–3.5 and Supplementary Table S4), Mann–Whitney U comparisons (n = 4; Section 3.5 and Supplementary Table S6), Kruskal–Wallis tests by age stratum (n = 3; Section 3.4), and Pearson's χ² (n = 1; Supplementary Table S6), and an autocorrelation test of the signed interocular difference against a measurement-error null (n = 1; Section 3.3). Clustering-robust re-estimations of associations already included in this inventory (within- and between-patient decompositions and cluster bootstraps in Sections 3.3 and 3.4), and partial correlations with additional covariate adjustment reported alongside a principal estimate, are re-analyses of the same hypotheses rather than independent tests. Sensitivity correlations in Section 3.6 and Supplementary Tables S1–S3 quantify agreement.

*Bootstrap confidence intervals.* Bootstrap 95 % confidence intervals for stratum-specific medians of CRAI and AAI (Table 3 legend) were computed by percentile method with 10,000 resamples of the within-stratum patient-level data, drawn with replacement.

*Robustness to RAI = 100 % boundary intervals.* Of 913 valid intervals, 44 (4.8 %) reached the boundary RAI = 100 %, comprising opposite-direction inter-eye changes (33/44 = 75 %; one eye elongating, the other shortening, typically reflecting asymmetric choroidal response to therapy) and one-eye-stable cases (11/44 = 25 %; one eye unchanged within measurement noise, the other progressing). Distribution by age stratum was non-monotonic (5–8 yr 2.3 %; 8–10 yr 5.5 %; 10–12 yr 7.7 %; 12–16 yr 2.7 %), with the lowest frequency in the oldest stratum, inconsistent with a clipping-driven explanation for the age-related rise of CRAI. Recomputation of patient-level CRAI excluding RAI = 100 % intervals (lost 3 patients with all intervals at the boundary; n = 264 retained) preserved the principal age association: Spearman ρ(CRAI, age) = +0.243, p = 6.7 × 10⁻⁵, compared with ρ = +0.275, p = 5.1 × 10⁻⁶ in the full specification.

*Variance decomposition of RAI and patient-level reliability.* Two complementary statistical quantities are reported. (i) The interval-level descriptive variance decomposition reported in Section 3.3 (~42 % between-patient, ~58 % within-patient) was computed by classical analysis-of-variance partitioning (between-patient sum of squares weighted by within-patient sample size), and answers the descriptive question of what fraction of RAI variability across all 913 intervals separates patient means from each other. (ii) The patient-level reliability of CRAI as an estimator of a patient's underlying asymmetric tendency was estimated by a random-intercept linear mixed model (REML, statsmodels MixedLM; RAI ~ 1 + (1 | patient)), yielding an Intraclass Correlation Coefficient (ICC) = 0.180 (variance between = 127.3, variance within = 578.3). The two quantities differ because the analytical cohort is unbalanced (71 patients with 1 valid interval, 196 with ≥ 2 intervals; median 3, IQR 1–5 intervals/patient); the ANOVA-style decomposition counts the full deviation of each 1-interval patient as "between" without the within-patient counterpart that would normally be discounted, inflating the descriptive between-patient share. The REML ICC is the proper reliability estimate for unbalanced designs. By the criteria of Koo and Li (2016), an ICC of 0.18 falls in the "poor" range, indicating that patient-level CRAI is not a stable individual trait.^[21]^ A sensitivity analysis restricting to patients with ≥ 2 valid intervals (n = 196, 842 intervals) confirmed the robustness of cohort-level findings: median CRAI 22.7 % (unchanged), Pronounced prevalence 13.8 % (vs 13.9 %), ρ(CRAI, age) = +0.312, p < 10⁻⁵ (vs +0.275 in the full cohort); the REML ICC was 0.184 (essentially identical), confirming that the reliability estimate is not driven by the inclusion of single-interval patients. Patients contributing a single valid interval (n = 71) showed a CRAI distribution indistinguishable from those with ≥ 2 intervals (median 22.7 % in both; pronounced asymmetry 15.5 % vs 13.8 %), and contributed 71 of 913 intervals (7.8 %).

***Autocorrelation test against a measurement-error null.*** Consecutive intervals of a patient share a visit: the measurement error at that visit enters the earlier interval's ΔAL with a positive sign and the later interval's with a negative sign. Under a null of symmetric bilateral elongation in which the entire signed interocular difference (ΔAL_OD − ΔAL_OS) arises from measurement error, this shared-visit structure implies a lag-1 autocorrelation of −0.5 across consecutive intervals, independent of the assumed instrumental SEM. Simulation confirmed this: symmetric truth with visit-level errors (diurnal component shared between fellow eyes, instrumental component independent) yielded lag-1 autocorrelation of −0.44 to −0.48 across σ_inst from 0.014 to 0.050 mm. The observed value in patients with ≥ 2 valid intervals was −0.18 (p = 5.9 × 10⁻⁶), substantially attenuated relative to the null. Because the null prediction is invariant to σ_inst, this attenuation cannot be reconciled with measurement error at any noise level, and implies a genuine asymmetric component with variance approximately twice that of measurement error in the interval-level interocular difference.

***Estimation of interocular measurement error.*** Let d_j_ denote the signed interocular AL difference (OD − OS) at visit j, equal to the true difference plus a visit-level measurement error e_j_ with SD σ_e_, and let δ_i_ = d_i+1_ − d_i_. Consecutive intervals share a visit, so Cov(δ_i_, δ_i+1_) = −σ_e_², provided that the biological increments of the interocular difference are serially independent and independent of measurement error. Non-adjacent intervals share no visit, so under the same assumptions corr(δ_i_, δ_i+2_) = 0; the lag-2 autocorrelation therefore serves as a check of serial independence, although it cannot exclude a process correlated at lag 1 only. Positive serial correlation of biological increments, as would arise from persistence of the leading eye, makes the lag-1 covariance less negative and biases σ_e_ downward; serially alternating increments have the opposite effect.

Because excluding intervals would break the shared-visit structure and the |ΔAL| filter would preferentially retain large changes, the estimate used all 1,121 raw consecutive visit pairs of the 267 analysed patients within the 5–16-year window (the 1,128 pairs in Supplementary Figure S1 additionally include one pair from each of seven patients excluded for interval-level filtering failure; single pairs carry no adjacent-pair information), without the interval-level filters of Section 2.6 and without trimming; the largest changes of the interocular difference (≤ 0.56 mm) were verified against source records. The lag-1 autocorrelation was −0.20 (854 adjacent pairs) and the lag-2 autocorrelation +0.04 (643 pairs). The resulting estimate was σ_e_ = 0.030 mm (patient-level cluster-bootstrap 95 % CI 0.015–0.044), i.e. 0.043 mm per interval; restricted to the 913 valid intervals, the same estimator gave 0.031 mm. Allowing for the small positive lag-2 covariance, under the assumption that biological covariance is similar at lags 1 and 2 (σ_e_² = γ₂ − γ₁, where γ_k_ is the lag-k autocovariance), gave 0.033 mm (95 % CI 0.017–0.045).

On intervals shorter than 120 days, where genuine growth is smallest, the SD of the change in the interocular difference was 0.58 times the single-eye SD, against 1.41 expected for independent errors, implying a correlation of approximately 0.8 between fellow eyes in short-term AL variability. Most visit-level variability is therefore shared between fellow eyes (plausibly diurnal choroidal variation, although part of the instrumental error may also be common) and cancels in the interocular difference. The single-eye measurement SD cannot be recovered by this estimator, because positively autocorrelated growth offsets the shared-visit term; it would require test–retest data.

The estimate increased with the combined duration of the paired intervals (0.019, 0.026 and 0.041 mm across duration tertiles), which pure measurement error would not do, whereas interocular differences did not converge towards zero over follow-up. The pooled estimate therefore probably includes a duration-dependent biological component and is likely conservative; the short-interval estimate is compatible with the lower end of reported within-session repeatability (Sw ≈ 0.014 mm,^[13]^ i.e. ≈ 0.020 mm for the interocular difference).

Because the confidence interval of σ_e_ is wide, the noise floor and MDC₉₅ were recomputed at its bounds, at the lag-2-adjusted estimate, at the short-interval estimate, and at the previously adopted value (Supplementary Table S7). The proportion of patients above their MDC₉₅ ranged from 10 % to 52 % (39 % at the short-interval estimate), and the majority of the pronounced-asymmetry stratum exceeded MDC₉₅ at every value (51–92 %). No pronounced-stratum patient fell below the noise floor at any value except the upper bound (one patient), and detectability did not decline with age at any value except the lower bound.

*Discrimination of the scale-invariance test.* Because RAI normalises the interocular difference by max(ΔAL_OD, ΔAL_OS) while the net AL change rate is that same quantity divided by interval duration, the two share a term, raising the concern that their near-zero partial correlation is structurally predetermined. This was tested by simulation. Two generative models were calibrated to reproduce the observed median CRAI of 22.7 %: (a) an asymmetric component proportional to total elongation, and (b) an asymmetric component of fixed absolute magnitude independent of elongation. Model (a) yielded partial ρ(CRAI, net rate | age) = +0.08 (95 % range −0.01 to +0.15); model (b) yielded −0.27 (−0.37 to −0.17). The observed value of −0.08 lies between these regimes, excluding both strict proportionality and elongation-independent asymmetry, and supporting approximate but sublinear scaling. Patient-level aggregation by the median across intervals breaks the algebraic dependence that would otherwise link the two quantities within a single interval.

*Annualisation artefact assessment.* Because AAI = |ΔAL_OD − ΔAL_OS| / duration_years, short intervals can inflate annualised estimates. Within the analytical cohort, median interval duration was 182 days (IQR 128–221), and AAI was negatively associated with duration (Spearman ρ = −0.26, p < 10⁻¹⁵), confirming this concern in principle. However, decomposition by AAI quartile showed that the raw un-annualised absolute interocular difference |ΔAL_OD − ΔAL_OS| also rose monotonically across quartiles (Q1 median 0.010 mm; Q4 median 0.090 mm — a 9-fold increase), indicating that the AAI top quartile reflects true high asymmetry rather than short-duration inflation alone. Interval-duration distributions were comparable across age strata (median 170–189 days; % intervals ≤ 180 days, 39–55 %), ruling out duration distribution as a driver of the age-related rise in AAI. As an independent specification check, a cumulative patient-level AAI computed as Σ|ΔAL_OD − ΔAL_OS| / Σ duration_years over each patient's full follow-up (n = 196 patients with ≥ 2 valid intervals, free of per-interval annualisation) yielded a median of 0.076 mm/year and preserved the positive age association (Spearman ρ = +0.23, p < 0.01), confirming that the age-related rise in AAI is not an artefact of annualisation. The 180-day sensitivity analysis (Section 3.6 and Supplementary Tables S1–S3) provides an additional check.

*Choice of age variable and follow-up confounding.* The age gradient of CRAI was examined against three age definitions: age at first visit (ρ = +0.275), at the midpoint of follow-up (+0.208), and at the last visit (+0.127). The attenuation is not attributable to age-related stabilisation of progression, which shows the opposite pattern (net AL change rate vs age at first visit ρ = −0.232; vs age at last visit ρ = −0.318), nor to therapy exposure, which is unrelated to CRAI (ρ = +0.094, p = 0.13; treated 22.2 % vs untreated 23.8 %, p = 0.71) and does not mediate the follow-up association (partial ρ unchanged at −0.133). It reflects instead the construction age_last = age_first + follow-up duration, combined with a negative association between CRAI and follow-up duration (ρ = −0.133, p = 0.03). The latter is an aggregation effect: CRAI is a median across intervals, and its dispersion contracts as intervals accumulate (interquartile width 31.3 percentage points at one interval vs 19.1 at ≥ 4). Adjustment for follow-up duration in addition to net AL change rate attenuated the age association from partial ρ = +0.252 to +0.216 (p = 0.0004); further adjustment for therapy exposure gave +0.205 (p = 0.0008). The interval-level within-patient analysis, unaffected by this construction, confirmed the gradient independently (r = +0.10, p = 0.006).

#### Supplementary Tables

**Supplementary Table S1.** Sensitivity to noise threshold (interval-level filter on max |ΔAL|).

| Threshold (mm) | n intervals | n patients | Median CRAI (%) | Median AAI (mm/year) | Pronounced asymmetry (%) | ρ vs principal |
| --- | --- | --- | --- | --- | --- | --- |
| 0.05 (principal) | 913 | 267 | 22.7 | 0.073 | 13.9 | — |
| 0.07 | 811 | 259 | 22.2 | 0.072 | 12.4 | 0.952 |
| 0.10 | 657 | 244 | 21.3 | 0.080 | 13.5 | 0.899 |

Variation in the noise threshold across a clinically plausible range preserves patient-level CRAI ranking (ρ ≥ 0.90) and prevalence estimates for pronounced asymmetry within ~2 percentage point of the principal specification.

**Supplementary Table S2.** Sensitivity to RAI aggregator and denominator definition (n = 267 patients).

*(a) Aggregator: function used to combine within-patient RAI values into CRAI.*

| Aggregator | Median CRAI (%) | IQR | ρ vs median | Concordant categorisation (%) |
| --- | --- | --- | --- | --- |
| Median (principal) | 22.7 | [12.8–38.8] | 1.000 | 100.0 |
| Arithmetic mean | 25.8 | [16.7–40.5] | 0.931 | 88.4 |
| 10 %-trimmed mean | 25.6 | [16.4–40.5] | 0.932 | 88.4 |

*(b) Denominator: choice of D in eq. (1) for Case 1 (both ΔAL non-negative).*

| Denominator D | Median CRAI (%) | ρ vs principal |
| --- | --- | --- |
| max (ΔAL_OD, ΔAL_OS) (principal) | 22.7 | 1.000 |
| Σ \|ΔAL\| | 13.4 | 0.991 |
| Mean of \|ΔAL\| | 25.8 | 0.997 |

Aggregator choice has a modest effect on the central tendency of CRAI but preserves patient ranking and category assignment in ≥ 88 % of patients. Denominator choice rescales CRAI values but preserves patient ranking essentially perfectly (ρ ≥ 0.99), confirming that the principal “max” specification reflects a methodological convention rather than a result-driving choice.

**Supplementary Table S3.** Sensitivity to interval duration filter.

| Filter | n intervals | n patients | Median CRAI (%) | Pronounced asymmetry (%) | Partial ρ for CRAI ↔ age, controlling for net AL change rate |
| --- | --- | --- | --- | --- | --- |
| ≥ 90 days (principal) | 913 | 267 | 22.7 | 13.9 | +0.25, p < 0.001 |
| ≥ 180 days | 481 | 206 | 20.2 | 14.6 | +0.13, p = 0.07 |

Spearman correlation of patient-level CRAI between the 90- and 180-day specifications (in the overlapping 206-patient subset) was ρ = 0.80. The prevalence estimate for pronounced asymmetry remained essentially unchanged (13.9 % vs 14.6 %). The age-related rise of CRAI was attenuated to marginally non-significant within the smaller 180-day cohort, consistent with reduced statistical power rather than a substantive change in the underlying pattern.

**Supplementary Table S4. Characterisation of self-normalisation:** RAI versus AAI in relation to absolute amplitude of axial change, decomposed by analytic level.

| Analytic level | Variable pair | Spearman ρ |
| --- | --- | --- |
| **Interval-level pooled (n = 913)** | RAI ↔ max growth | −0.23 |
|  | AAI (raw) ↔ max growth | +0.33 |
| **Within-patient (mean-centred)** | RAI ↔ max growth | −0.27 |
|  | AAI (raw) ↔ max growth | +0.29 |
| **Between-patient (patient means)** | RAI ↔ max growth | −0.18 |
|  | AAI (raw) ↔ max growth | +0.37 |
| **Patient-level, bivariate (n = 267)** | CRAI ↔ annualised net AL change rate | −0.14, p = 0.025 |
|  | AAI (annualised) ↔ rate | +0.28, p < 0.001 |
| **Patient-level, partial (age controlled)** | CRAI ↔ rate \| age | −0.08, p = 0.20 |
|  | AAI (annualised) ↔ rate \| age | +0.35, p < 0.001 |

Across all analytic levels, RAI shows weak negative or null associations with the absolute amplitude of axial change, whereas AAI shows consistently positive associations. The dissociation confirms the methodological design intent: RAI captures the relative pattern of asymmetry, scale-invariant to the overall amplitude of progression, while AAI quantifies the absolute rate of interocular discordance. At the patient level, after adjustment for age — itself associated with net AL change rate — CRAI is statistically independent of the overall pace of net AL change.

**Supplementary Table S5.** Comparison of cut-off systems for CRAI categorisation (n = 267 patients).

| Cut-offs | Category | n | % (95 % CI) | Median AAI (mm/year) | % below noise (0.057) | % above patient-specific MDC₉₅ |
| --- | --- | --- | --- | --- | --- | --- |
| **10 / 30 %** | Minimal (≤ 10) | 44 | 16.5 (12.5–21.4) | 0.020 | 91 | 0 |
|  | Moderate (11–30) | 125 | 46.8 (40.9–52.8) | 0.059 | 48 | 6 |
|  | Pronounced (> 30) | 98 | 36.7 (31.1–42.6) | 0.121 | 5 | 43 |
| **15 / 45 %** | Minimal (≤ 15) | 82 | 30.7 (25.5–36.5) | 0.026 | 88 | 1 |
|  | Moderate (16–45) | 135 | 50.6 (44.6–56.5) | 0.083 | 24 | 13 |
|  | Pronounced (> 45) | 50 | 18.7 (14.5–23.8) | 0.158 | 0 | 62 |
| **15 / 50 %** (principal) | Minimal (≤ 15) | 82 | 30.7 (25.5–36.5) | 0.026 | 88 | 1 |
|  | Moderate (16–50) | 148 | 55.4 (49.4–61.3) | 0.085 | 22 | 15 |
|  | Pronounced (> 50) | 37 | 13.9 (10.2–18.5) | 0.218 | **0** | 70 |

The principal 15 / 50 % specification was chosen on principled grounds: (i) the 15 % threshold is a statistical noise filter, with 88 % of patients in this stratum having AAI below the patient-level measurement noise floor (~0.057 mm/year); (ii) the 50 % threshold has a direct biological interpretation, corresponding to the slower-progressing eye accumulating axial length at no more than half the rate of the faster-progressing eye over the median interval. The Pronounced stratum under this scheme provides the strongest convergent support against measurement uncertainty: no patients fall below the noise floor, and 70 % exceed their patient-specific MDC₉₅. The convention-based 10 / 30 % scheme yields a larger Pronounced stratum (37 %) with substantially lower specificity (43 % above MDC₉₅; 5 % below noise) and is reported for comparability with categorical schemes commonly used in adult anisometropia literature. The 15 / 45 % alternative produces intermediate results.

**Supplementary Table S6.** Post-hoc therapy-stratified description of CRAI and AAI in the analytical cohort (n = 267). Primary therapy per patient was defined as the category of the therapy episode of greatest duration during follow-up; ten source categories were grouped into six analytical groups (see Supplementary Methods). The orthokeratology stratum is reported separately because of its potential for transient axial-biometry effects through anterior-segment changes. Wilson 95 % confidence intervals are reported for pronounced-asymmetry prevalence.

| **Therapy group** | **n (% of cohort)** | **Median CRAI, %** | **Median AAI, mm/year** | **n Pronounced (% of group)** | **95 % Wilson CI for Pronounced %** |
| --- | --- | --- | --- | --- | --- |
| No active therapy | 122 (45.7) | 19.7 | 0.072 | 9 (7.4) | 3.9–13.4 |
| Atropine monotherapy | 61 (22.8) | 28.6 | 0.080 | 12 (19.7) | 11.6–31.3 |
| Atropine + non-OK optical | 15 (5.6) | 33.3 | 0.080 | 4 (26.7) | 10.9–52.0 |
| Orthokeratology (any) | 55 (20.6) | 23.8 | 0.074 | 11 (20.0) | 11.6–32.4 |
| Optical monotherapy (non-OK) | 13 (4.9) | 21.0 | 0.049 | 1 (7.7) | 1.4–33.3 |
| Other / multi-optical | 1 (0.4) | 11.1 | 0.024 | 0 (0.0) | 0.0–79.3 |
| **Overall** | **267 (100.0)** | **22.7** | **0.073** | **37 (13.9)** | **10.2–18.5** |

Pearson's χ² test of CRAI category (Minimal / Moderate / Pronounced) by six therapy groups: χ² = 15.5, dof = 10, p = 0.11 (not significant). Mann–Whitney U comparisons between patients with orthokeratology as primary therapy (n = 55) and all other patients (n = 212): CRAI p = 0.78; AAI p = 0.97. A sensitivity analysis excluding all orthokeratology patients yielded pronounced-asymmetry prevalence of 12.3 % (vs 13.9 % overall) — a modest shift consistent with the absence of OK-specific confounding. Within the no-therapy subset alone (n = 122), pronounced asymmetry was observed in 7.4 % of patients (95 % Wilson CI 3.9–13.4), demonstrating that the phenomenon is present in patients with no concurrent treatment artefacts.

Source categories grouped into each analytical group: Group 1 = no_therapy. Group 2 = atropine_mono. Group 3 = atropine_plus_soft, atropine_plus_defocus_glasses. Group 4 = ortho_mono, atropine_plus_ortho. Group 5 = soft_lens_mono, defocus_glasses_mono. Group 6 = multi_optical, other_combination.

**Supplementary Table S7.** Sensitivity of the noise floor, MDC₉₅ and detectability to the estimate of interocular measurement error σ_e_ (n = 267 patients).

| **σ_e_, mm** | **Basis** | **Noise floor*** | **Median MDC₉₅*** | **Above MDC₉₅, n (%)** | **Below noise floor, %: all / Minimal** | **Pronounced: above MDC₉₅, % / below floor, n** | **Above MDC₉₅ by age stratum, %†** |
| --- | --- | --- | --- | --- | --- | --- | --- |
| 0.015 | Lower 95 % CI bound | 0.028 | 0.069 | 139 (52.1) | 18 / 56 | 92 / 0 | 49.2 / 57.1 / 53.5 / 41.7 |
| 0.019 | Short-interval estimate | 0.035 | 0.085 | 105 (39.3) | 22 / 68 | 86 / 0 | 35.2 / 42.9 / 44.2 / 41.7 |
| **0.030** | **Principal estimate** | **0.057** | **0.138** | **49 (18.4)** | **39 / 88** | **70 / 0** | **14.8 / 20.2 / 20.9 / 33.3** |
| 0.033 | Lag-2-adjusted estimate | 0.062 | 0.150 | 43 (16.1) | 42 / 90 | 65 / 0 | 12.5 / 17.9 / 18.6 / 33.3 |
| 0.035 | Previously adopted value | 0.067 | 0.162 | 39 (14.6) | 45 / 91 | 65 / 0 | 11.7 / 15.5 / 16.3 / 33.3 |
| 0.044 | Upper 95 % CI bound | 0.083 | 0.202 | 26 (9.7) | 55 / 94 | 51 / 1 | 7.8 / 8.3 / 11.6 / 33.3 |

σ_e_, visit-level measurement-error SD of the signed interocular AL difference (Supplementary Methods, Estimation of interocular measurement error). All rows were computed by the same simulation (20,000 replicates per patient, fixed seed) with only σ_e_ varied; the principal row reproduces Table 2 and Section 3.4. The previously adopted value corresponds to a per-measurement SD of 0.025 mm per eye. Minimal and Pronounced refer to the principal CRAI categories (≤ 15 % and > 50 %). The short-interval estimate was obtained from the shortest tertile of paired-interval duration. *mm/year. †Age at first visit: 5–8 / 8–10 / 10–12 / 12–16 years (n = 128 / 84 / 43 / 12).

#### Supplementary Figures

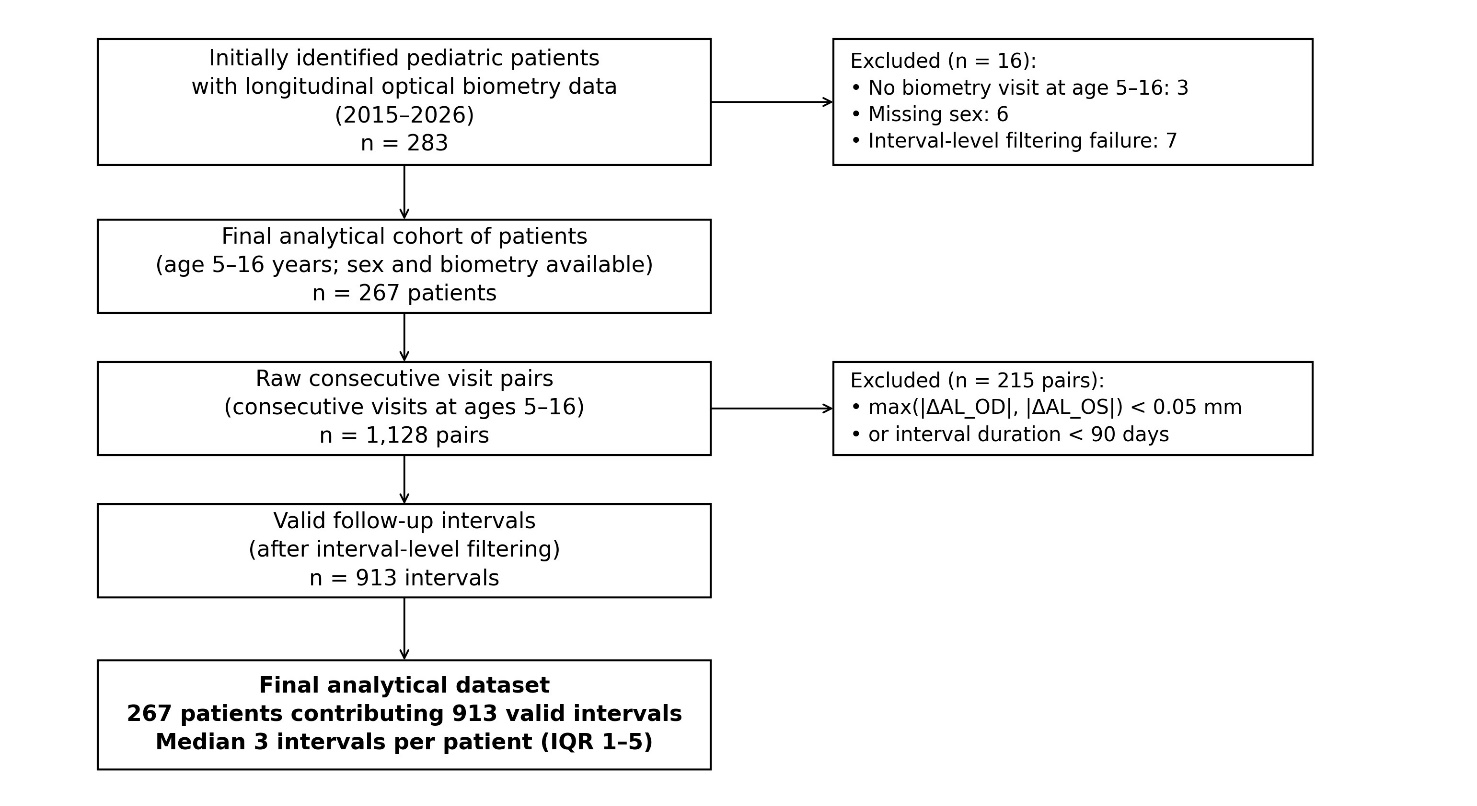

**Supplementary Figure S1.** *STROBE flow diagram of patient and interval selection.* Of 283 initially identified paediatric patients with longitudinal optical biometry data collected during 2015–2026, 16 were excluded (3 with no biometry visit at age 5–16, 6 with missing sex, 7 failing interval-level filtering criteria), yielding a final analytical cohort of 267 patients. Within the eligible cohort, 1,128 raw consecutive visit pairs (consecutive visits at ages 5–16) were generated; application of the interval-level filters (max(|ΔAL_OD|, |ΔAL_OS|) ≥ 0.05 mm; duration ≥ 90 days) yielded 913 valid follow-up intervals contributing to the analysis.
